# Real-World Effectiveness of Casirivimab and Imdevimab Among Patients Diagnosed With COVID-19 in the Ambulatory Setting: A Retrospective Cohort Study Using a Large Claims Database

**DOI:** 10.1101/2022.05.19.22272842

**Authors:** Mohamed Hussein, Wenhui Wei, Vera Mastey, Robert J. Sanchez, Degang Wang, Dana J. Murdock, Boaz Hirshberg, David M. Weinreich, Jessica J. Jalbert

**Affiliations:** Regeneron Pharmaceuticals, Inc., Tarrytown, New York

## Abstract

**Importance:** Data on real-world effectiveness of casirivimab and imdevimab (CAS+IMD) for treatment of coronavirus 2019 (COVID-19) are limited, especially with regard to variants of concern such as Delta.

**Objective:** To assess effectiveness of CAS+IMD versus no COVID-19 antibody treatment among patients diagnosed with COVID-19 in the ambulatory setting overall and among subgroups, including patients diagnosed during the Delta-dominant period prior to Omicron emergence.

**Design:** Retrospective cohort study.

**Setting:** Komodo Health closed claims database.

**Participants:** Patients diagnosed with COVID-19 in ambulatory settings from December 2020 through September 2021 treated with CAS+IMD or untreated but treatment-eligible under the Emergency Use Authorization (EUA). Each treated patient was exact- and propensity score-matched without replacement to up to 5 untreated EUA-eligible patients.

**Exposure:** CAS+IMD treatment.

**Main Outcomes and Measures:** Composite endpoint of 30-day all-cause mortality or COVID-19-related hospitalization. Kaplan-Meier estimators were used to calculate risk of outcome overall and across subgroups defined by age groups, COVID-19 vaccination status, immunocompromised, and timing of COVID-19 diagnosis (December 2020 to June 2021, and July 2021 to September 2021). Cox proportional-hazards models were used to estimate adjusted hazard ratios (aHR) and 95% confidence intervals (95% CI).

**Results:** Among 75 159 patients treated with CAS+IMD and 1 670 338 EUA-eligible untreated patients, 73 759 treated patients were matched to 310 688 untreated patients; matched patients had an average age ∼50 years, approximately 60% were women and were generally well-balanced across risk factors. The 30-day risk of the composite outcome was 2.1% and 5.2% in the CAS+IMD -treated and untreated patients, respectively; CAS+IMD treatment was associated with a 60% lower risk of the outcome (aHR 0.40; 95% CI, 0.38-0.42). The effect of CAS+IMD treatment was consistent across subgroups, including those who received a COVID-19 vaccine (aHR 0.48, 95% CI, 0.41-0.56), and those diagnosed during the Delta-dominant period (aHR 0.40, 95% CI, 0.38-0.42).

**Conclusions and Relevance:** The real-world effectiveness of CAS+IMD is consistent with the efficacy for reducing all-cause mortality or COVID-19-related hospitalization reported in clinical trials. Effectiveness is maintained across patient subgroups, including those who may be prone to breakthrough infections, and was effective against susceptible variants including Delta.□

## INTRODUCTION

Clinical studies have identified neutralizing monoclonal antibodies (mAbs) as efficacious agents of passive immunotherapy for early treatment of patients diagnosed with COVID-19 in the ambulatory setting.^1-4^ These mAbs, which include casirivimab and imdevimab (CAS+IMD; Regeneron, Inc.), bamlanivimab with etesevimab (Eli Lilly), and sotrovimab (GlaxoSmithKline), have received Emergency Use Authorization (EUA) from the US Food and Drug Administration for treatment of non-hospitalized patients ≥12 years old who have mild-to-moderate COVID-19 and are at high risk of progressing to severe disease, including hospitalization or death.^5-7^

While mAbs demonstrated efficacy in COVID-19 patients, emergence of additional variants of concern (VOC) after the completion of clinical trials highlights the need for continued evaluation of mAbs in the real world. Delta (B.1.617.2) and Delta plus (AY.4.2) are VOC that are associated with higher infection rates^8,9^ and increased hospitalization and mortality.^10-12^ *In vitro* studies suggested that CAS+IMD retains neutralization activity against most VOC including Delta but with the exception of Omicron (B.1.1.529).^5,13-17^ Most real-world studies assessing the effectiveness of CAS+IMD for treating COVID-19^18-30^ were conducted prior to Delta, and those that did overlap with the emergence of the Delta variant were single-center and of small sample size.^19,30,31^ Furthermore, with the introduction of COVID-19 vaccines, an improved understanding of vulnerable groups, and the emergence of the Delta variant, it became important to evaluate the effectiveness of CAS+IMD across these patient subgroups. Therefore, the objective of this study was to assess the effectiveness of CAS+IMD compared to no COVID-19 antibody treatment on 30-day all-cause mortality or COVID-19-related hospitalization among patients diagnosed with COVID-19 in the ambulatory setting overall and among patient subgroups, including among patients diagnosed with COVID-19 during the Delta-dominant period.

## METHODS

### Study design and patient population

Based on a protocol in which we prespecified our analysis, we conducted a retrospective cohort study using the closed administrative claims data from the Komodo Health claims database; the closed claims dataset contains complete medical and prescription claims information. As of September 30, 2021, approximately 98 000 patients in the dataset who had a COVID-19 diagnosis (ICD-10-CM code U07.1) received intravenous or subcutaneous CAS+IMD. Since all data are de-identified and fully compliant with HIPAA, institutional review board/ethics committee approval was not required for this study.

Among patients diagnosed with COVID-19 in the outpatient setting between December 1, 2020, and September 30, 2021, we identified patients treated with CAS+IMD and patients who were eligible to be treated with CAS+IMD under the EUA but were untreated. Among treated patients, the index date was the date of CAS+IMD administration. For untreated patients, the index date was an assigned date matching the distribution of days from COVID-19 diagnosis to treatment for the CAS+IMD-treated patients. Additional inclusion criteria were continuous enrollment for ≥6 months pre-index (ie, baseline); age ≥12 years at index; a COVID-19 diagnosis within 10 days prior to (days 0 to –10) or on the index (day 0) but no diagnosis in the prior 30-day period (days –11 to –40); and a valid date of death. Patients were excluded if they were treated with other COVID-19 mAbs over the baseline period or on the index date or received CAS+IMD during baseline.

### Outcomes

The study outcome was the composite of 30-day all-cause mortality or COVID-19-related hospitalization. Sources used by Komodo to identify mortality included Social Security Administration (SSA) data, a private obituary data source, and a private claims mortality dataset. COVID-19-related-hospitalization was defined as a COVID-19 diagnosis as the primary or admitting diagnosis. Patients were followed from the index date until the occurrence of the outcome or a censoring event, which included receipt of another COVID-19 mAb, the end of 30-day risk period, healthcare plan disenrollment, or study end date (September 30, 2021).

### Study variables

Baseline demographic variables included age (as a continuous variable and categorized as 12-17, 18-54, 55-64, ≥65 years), sex, and geographic region. COVID-19-related variables included location of diagnosis (emergency room/urgent care vs other), the number of days from diagnosis to index date, the index month, and vaccinated against COVID-19 (ie, receipt ≥1 dose). The Charlson Comorbidity Index (CCI) score was derived using the presence of comorbidities over the baseline period.^32^ Body mass index (BMI) was categorized as normal, overweight, and obese based on ICD-10-CM diagnosis codes over the baseline period (including index date). The occurrence of ≥1 all-cause hospitalization and ≥1 all-cause emergency room/urgent care visit during the baseline period was also captured. Specific risk factors for use of CAS+IMD under the EUA were identified during the baseline period. These risk factors included age ≥65 years on index date; age 12-17 years on index date with BMI ≥85^th^ percentile for age and sex based on CDC growth charts^33^; BMI >25 kg/m^2^; pregnancy; diabetes; chronic lung disease; immunosuppressive disease; history of immunosuppressive treatment; cardiovascular disease, hypertension, or congenital heart disease; sickle cell disease; and neurodevelopmental disorders. The following EUA risk factors were evaluated over the baseline period only and did not include the index date since COVID-19 could result in these conditions: chronic kidney disease, cardiovascular disease or hypertension, and use of medical-related technological dependence.

### Subgroups

Subgroups of interest included age groups of 12-17, 18-54, 55-64, and ≥65 years; elevated risk defined as age ≥65 years or 55-64 years with BMI ≥35 kg/m^2^, type 2 diabetes, chronic obstructive pulmonary disease, or chronic kidney disease; immunocompromised, ie, B-cell deficiencies, both overall and by type of deficiency (primary, secondary, and drug-induced as defined in Supplementary Table 1; vaccinated against COVID-19; and timing of COVID-19 diagnosis (December 2020 to June 2021 vs July 2021 onward, which is the month the Delta variant became the dominant variant in the US).^34^ A post-hoc analysis was also conducted among vaccinated patients who were also at elevated risk based on the above definition.

### Statistical analysis

#### Matching

Propensity scores (PS), derived using logistic regression, predicted the probability of CAS+IMD treatment vs no treatment given age, sex, index month, 3-digit zip code, days between COVID-19 diagnosis and index date, individual EUA criteria, BMI category, CCI score, COVID-19 vaccination status, and baseline health care resource utilization. CAS+IMD -treated patients were then matched using a caliper of 0.2 of the standard deviation of the logit of the estimated PS^35^ and exact matched to up to 5 untreated EUA-eligible patients without replacement on the index month, 3-digit zip code, and days from COVID-19 diagnosis to index date. Missing data were accounted for by including an indicator variable. Balance between groups was measured by standardized mean differences (SMD) with value ≥0.1 indicating imbalance.^36^

#### Primary analysis

Baseline characteristics including means, standard deviations, medians, and interquartile ranges (IQR) for continuous variables, and frequency and percent for categorical variables are reported among treated and untreated matched patients. Kaplan-Meier estimators were used to estimate the 30-day risk of the composite outcome among the matched patients,^37^ with 95% confidence bands across the entire KM survival curves constructed using the Hall-Wellner method.^38^ Log-ranks tests were used to compare survival distributions.

Adjusted hazard ratios (aHR) with 95% confidence intervals (CI) were derived using Cox proportional-hazards models that fit the model to the matched pairs, and used sandwich variance estimators to account for clustering within matched sets.^39^

#### Subgroup analyses

The 30-day outcome risk for each subgroup was derived using KM estimators. Cox proportional-hazards models were used to estimate the effectiveness of CAS+IMD across subgroups using interaction terms between treatment and the subgroup in the model; results are presented as the aHR with its 95% CI. These estimates were derived from the same matched set of patients as the primary analysis and accounted for clustering of patients without adjustment for multiplicity of testing.

#### Sensitivity analyses

Sensitivity analyses included modifying the definition of COVID-19-related hospitalization to having a COVID-19 diagnosis in the primary position, requiring that only the untreated patients meet EUA criteria, shortening the time window between diagnosis and treatment to 5 days from 10 days, and using 3 months pre-index continuous enrollment instead of 6 months.

The analytic file was created and all analyses were conducted using SAS software (version 9.4, SAS Institute Inc., Cary, NC).

## RESULTS

### Patient populations

Among 13 273 128 patients who had a COVID-19 diagnosis during the study period in the closed claims Komodo dataset, 75 159 patients who received CAS+IMD and 1 670 338 untreated patients met study criteria and were eligible for matching. Prior to matching, the groups were imbalanced on several variables (Supplementary Table 2).

Among treated patients, 73 759 were successfully matched to 310 688 patients who were EUA-eligible but untreated (Supplementary Figure 1). After matching, the SMDs indicated no imbalance between treated and matched EUA-eligible untreated patients on any of the baseline variables (Table 1). Treated and EUA-eligible untreated patients were primarily female (∼60%), with a mean age ∼50 years, with greatest representation from the South (65-67%). The mean (SD) and median (IQR) number of days from diagnosis to index date in the CAS+IMD-treated cohort were 1.6 (2.1) and 1 (3) days, respectively, and the timing was comparable for the assigned index dates for the EUA-eligible untreated patients. Among the individual EUA criteria, the combination of cardiovascular disease, hypertension, and congenital heart disease had the highest prevalence (53-55%) followed by neurodevelopmental disorders (∼37%) and being overweight (34-36%) (Table 1).

**Table 1.** Baseline Characteristics of the Matched Cohorts.

| **Variable** | **CAS+IMD**<br>**(n = 73 759)** | **EUA-eligible**<br>**untreated**<br>**(n = 310 688)** | **SMD<sup>a</sup>** |
| --- | --- | --- | --- |
| **Age, years** |  |  |  |
| Mean ± SD (range) | 49.8 ±15.5 | 50.0 ± 16.6 | 0.01 |
| Median (IQR) | 51 (22) | 51 (23) | — |
| Range | 12-89 | 12-89 | — |
| **Age group, years, n (%)** |  |  |  |
| 12-17 | 1651 (2.2) | 8515 (2.7) | 0.03 |
| 18-34 | 11 395 (15.5) | 50 994 (16.4) | 0.03 |
| 35-44 | 13 656 (18.5) | 56 873 (18.3) | 0.01 |
| 45-54 | 16 036 (21.7) | 64 951 (20.9) | 0.02 |
| 55-64 | 19 245 (26.1) | 73 742 (23.7) | 0.06 |
| 65-74 | 8156 (11.1) | 33 339 (10.7) | 0.01 |
| 75-84 | 2829 (3.8) | 15 727 (5.1) | 0.06 |
| ≥85 | 791 (1.1) | 6547 (2.1) | 0.08 |
| **Sex, n (%)** |  |  |  |
| Female | 43 989 (59.6) | 187 438 (60.3) | 0.01 |
| Male | 29 770 (40.4) | 123 250 (39.7) | 0.01 |
| **Region, n (%)** |  |  |  |
| Midwest | 11 969 (16.2) | 51 787 (16.7) | 0.01 |
| Northeast | 6914 (9.4) | 32 968 (10.6) | 0.04 |
| South | 49 744 (67.4) | 203 380 (65.5) | 0.04 |
| West | 5132 (7.0) | 22 553 (7.3) | 0.01 |
| BMI category, n (%)<sup>b</sup> |  |  |  |
| Not overweight | 2103 (2.9) | 9242 (3.0) | 0.01 |
| Overweight (25-<30 kg/m<sup>2</sup>) | 5991 (8.1) | 26 110 (8.4) | 0.01 |
| Obese (30-<35 kg/m<sup>2</sup>) | 6829 (9.3) | 28 835 (9.3) | 0.00 |
| Severely obese (35-<40 kg/m<sup>2</sup>) | 5721 (7.8) | 22 672 (7.3) | 0.02 |
| Morbidly obese (≥40 kg/m<sup>2</sup>) | 7851 (10.6) | 28 854 (9.3) | 0.05 |
| Missing | 45 264 (61.4) | 194 975 (62.8) | 0.03 |
| CCI score, mean ± SD | 1.18 ± 1.74 | 1.11 ± 1.74 | 0.04 |
| Hospitalization during baseline<br>period, n (%) | 10 154 (13.8) | 41 451 (13.3) | 0.01 |
| Month of index date, n (%) |  |  |  |
| December 2020 | 668 (0.9) | 3258 (1.1) | 0.02 |
| January 2021 | 1910 (2.6) | 9303 (3.0) | 0.03 |
| February 2021 | 984 (1.3) | 4538 (1.5) | 0.01 |
| March 2021 | 1015 (1.4) | 4697 (1.5) | 0.01 |
| April 2021 | 1345 (1.8) | 6067 (2.0) | 0.01 |
| May 2021 | 720 (1.0) | 2989 (1.0) | 0.00 |
| June 2021 | 474 (0.6) | 1823 (0.6) | 0.01 |
| July 2021 | 6374 (8.6) | 27 698 (8.9) | 0.01 |
| August 2021 | 31 190 (42.3) | 132 837 (42.8) | 0.01 |
| September 2021 | 29 079 (39.4) | 117 478 (37.8) | 0.03 |
| Time from diagnosis to index date,<br>days |  |  |  |
| Mean ± SD | 1.6 ± 2.1 | 1.5 ± 2.1 | 0.03 |
| Median (IQR) | 1 (3) | 1 (3) | — |
| Range | 0-10 | 0-10 | — |
| Time from diagnosis to index date,<br>days, n (%) |  |  |  |
| 0 | 33 694 (45.7) | 149 534 (48.1) | 0.05 |
| 1 | 12 442 (16.9) | 49 223 (15.8) | 0.03 |
| 2 | 8768 (11.9) | 34 176 (11.0) | 0.03 |
| ≥3 | 18 855 (25.6) | 77 755 (25.0) | 0.01 |
| Initial outpatient COVID-19<br>diagnosis in ER, n (%) | 22 075 (29.9) | 83 191 (26.8) | 0.07 |
| Vaccinated | 12 983 (17.6) | 52 681 (17.0) | 0.02 |
| EUA criteria |  |  |  |
| Age ≥65 years | 11 776 (16.0) | 55 613 (17.9) | 0.05 |
| Children overweight<sup>c</sup> | 607 (0.8) | 2199 (0.7) | 0.01 |
| Overweight | 26 392 (35.8) | 106 471 (34.3) | 0.03 |
| Pregnancy | 2653 (3.6) | 12 322 (4.0) | 0.02 |
| Chronic kidney disease | 3003 (4.1) | 12 237 (3.9) | 0.01 |
| Diabetes | 19 259 (26.1) | 74 881 (24.1) | 0.05 |
| Chronic pulmonary disease | 15 904 (21.6) | 63 419 (20.4) | 0.03 |
| Immunosuppressive disease | 8321 (11.3) | 31 189 (10.0) | 0.04 |
| Immunosuppressant use | 3428 (4.7) | 10 303 (3.3) | 0.07 |
| Sickle cell disease | 211 (0.3) | 913 (0.3) | 0.00 |
| Cardiovascular disease,<br>hypertension, or congenital heart<br>disease | 40 316 (54.7) | 163 107 (52.5) | 0.04 |
| Neurodevelopmental disorders | 27 579 (37.4) | 114 661 (36.9) | 0.01 |
| Medical-related technological<br>dependence | 18 840 (25.5) | 75 496 (24.3) | 0.03 |
| B-cell deficiency | 4483 (6.1) | 14 185 (4.6) | 0.07 |
| Primary | 15 (<0.1) | 33 (<0.1) | 0.01 |
| Secondary | 183 (0.3) | 560 (0.12) | 0.02 |
| Drug-induced | 4285 (5.8) | 13 592 (4.4) | 0.07 |
| Cancer or chemotherapy | 8486 (11.5) | 29 069 (9.4) | 0.07 |
| Cancer | 6199 (8.4) | 21 280 (6.9) | 0.06 |

| **Variable** | **CAS+IMD**<br>**(n = 73 759)** | **EUA-eligible**<br>**untreated**<br>**(n = 310 688)** | **SMD<sup>a</sup>** |
| --- | --- | --- | --- |
| Chemotherapy | 3650 (5.0) | 11 579 (3.7) | 0.06 |

Abbreviations: BMI, body mass index; CAS+IMD, casirivimab and imdevimab; CCI, Charlson Comorbidity Index; ER, emergency room; EUA, Emergency Use Authorization; IQR, interquartile range; SD, standard deviation; SMD, standardized mean difference.
<sup>a</sup> Value $\geq 0.1$ indicates significant imbalance between cohorts.
<sup>b</sup> Based on diagnoses relating to the BMI categories.
<sup>c</sup> Based on BMI $\geq 85$ th percentile for age and sex among those 12-17 years old.

### Primary analysis

The 30-day risk of all-cause mortality or COVID-19-related hospitalizations was 2.1% (95% CI, 2.0-2.2) in the CAS+IMD -treated cohort and 5.2% (95% CI, 5.1-5.3) in the EUA-eligible untreated cohort (Figure 1) representing 1486 and 15 027 events, respectively. Most of the events in both cohorts occurred within the first 10 days post-index. In adjusted models, CAS+IMD was associated with a 60% lower risk of the composite outcome compared to the untreated EUA eligible patients (aHR 0.40; 95% CI, 0.38-0.42) (Figure 2). The number of deaths observed during the 30-day post-index period was lower in the treated cohort relative to those who did not receive treatment, 51 and 1491, respectively, and the 30-day mortality risk was 0.1% (95% CI 0.06% to 0.11%) in the treated cohort and 0.6% (95% CI 0.56% to 0.62%) in the untreated cohort, respectively. Multiple sensitivity analyses showed results that were consistent with the primary analysis (Figure 2)

**Figure 1.**
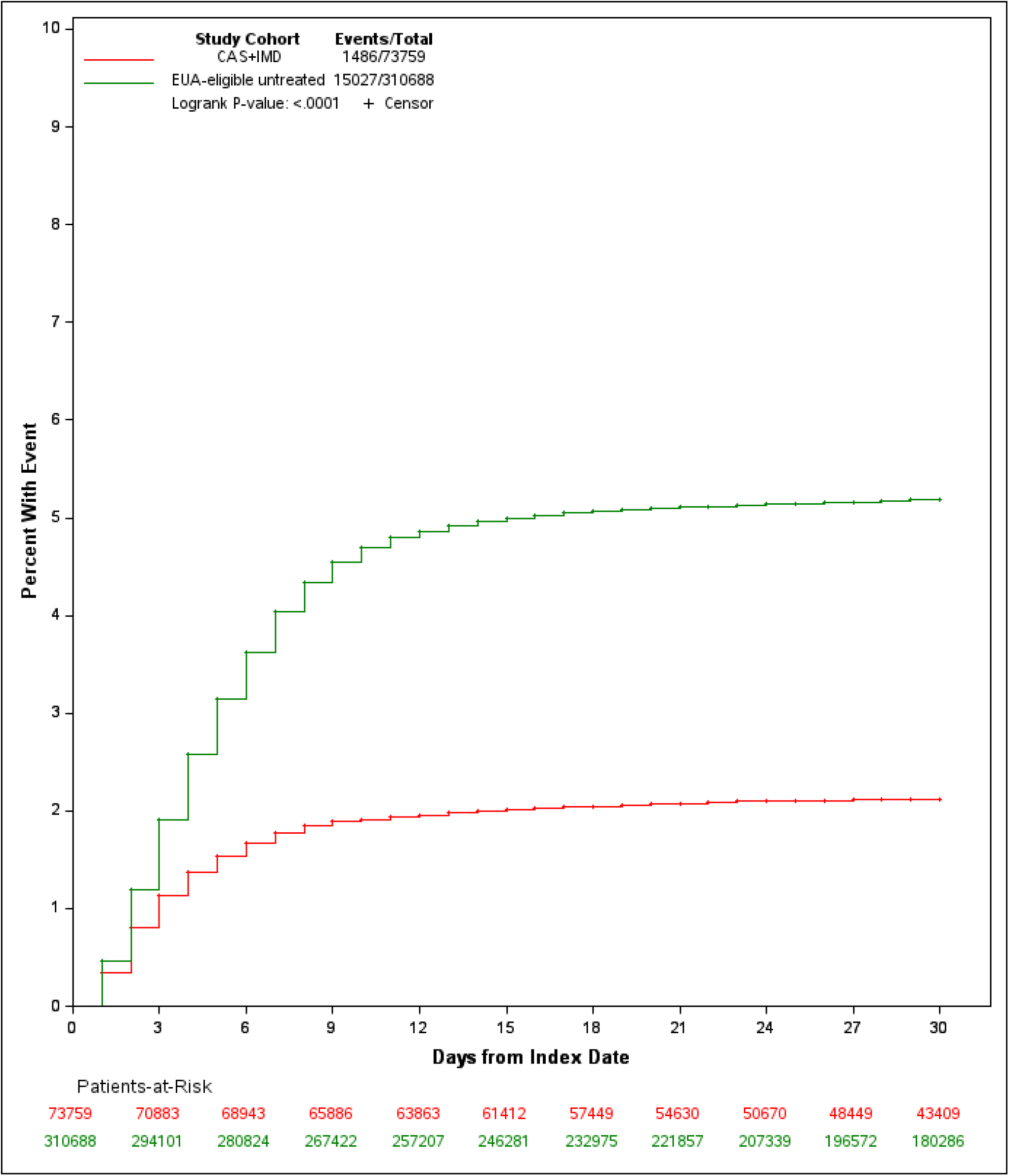
Kaplan-Meier Curve for 30-Day All-Cause Mortality or COVID-19–Related Hospitalization Among Patients Diagnosed With COVID-19 in the Outpatient Setting Who Were Treated With CAS+IMD or Who Were EUA-Eligible But Untreated.

**Figure 2.**
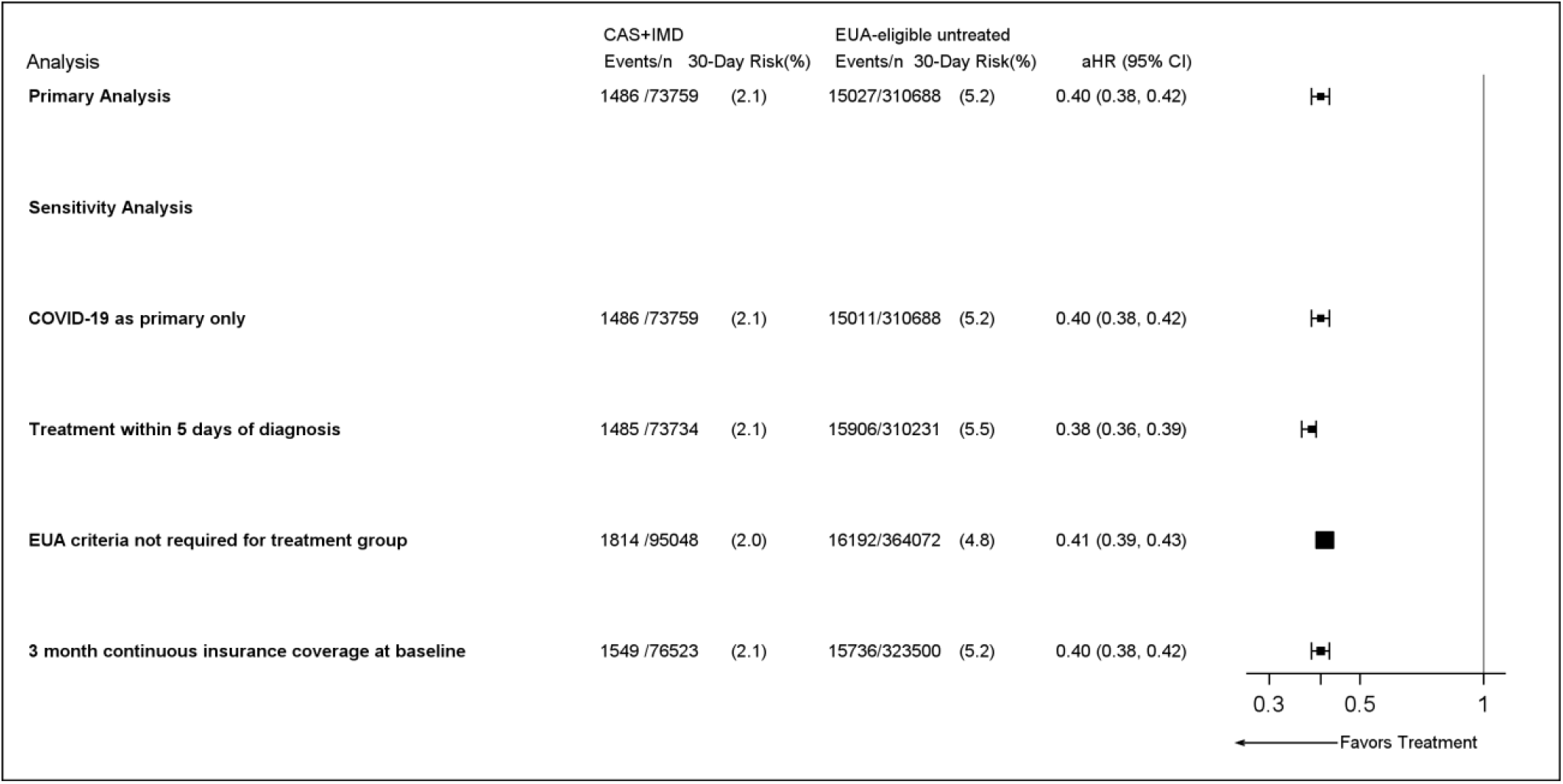
Primary and Sensitivity Analyses of 30-Day All-Cause Mortality or COVID-19– Related Hospitalization Among Patients Diagnosed With COVID-19 in the Outpatient Setting. Abbreviations: aHR, adjusted hazard ratio; CI, confidence interval.

### Subgroup analyses

The 30-day outcome risk among untreated EUA-eligible patients was highest for patients in the oldest age groups, and among those at elevated risk or who are immunocompromised (B-cell deficient) (Figure 4). After matching, the effectiveness of treatment with CAS+IMD was consistent across patient subgroups defined by age, COVID-19 vaccination status, elevated risk, and immunocompromised (Figure 4); there was a greater risk reduction among those with primary or secondary B-cell deficiency, although the numbers are small. Post-hoc analysis also showed that the treatment was associated with a 60% reduction in risk among vaccinated patients who were also at elevated risk (aHR 0.40; 95% CI 0.28–0.57).

Regardless of whether they were treated during the Delta-dominant period or not, patients who received CAS+IMD had a lower risk than EUA-eligible non-treated patients (Figure 3). Treatment with CAS+IMD was associated with a 50% lower risk (aHR 0.50; 95% CI, 0.43-0.58) during the earlier period, and a 60% lower risk (aHR 0.40; 95% CI, 0.38-0.42) during Delta-dominant period (Figure 4).

**Figure 3.**
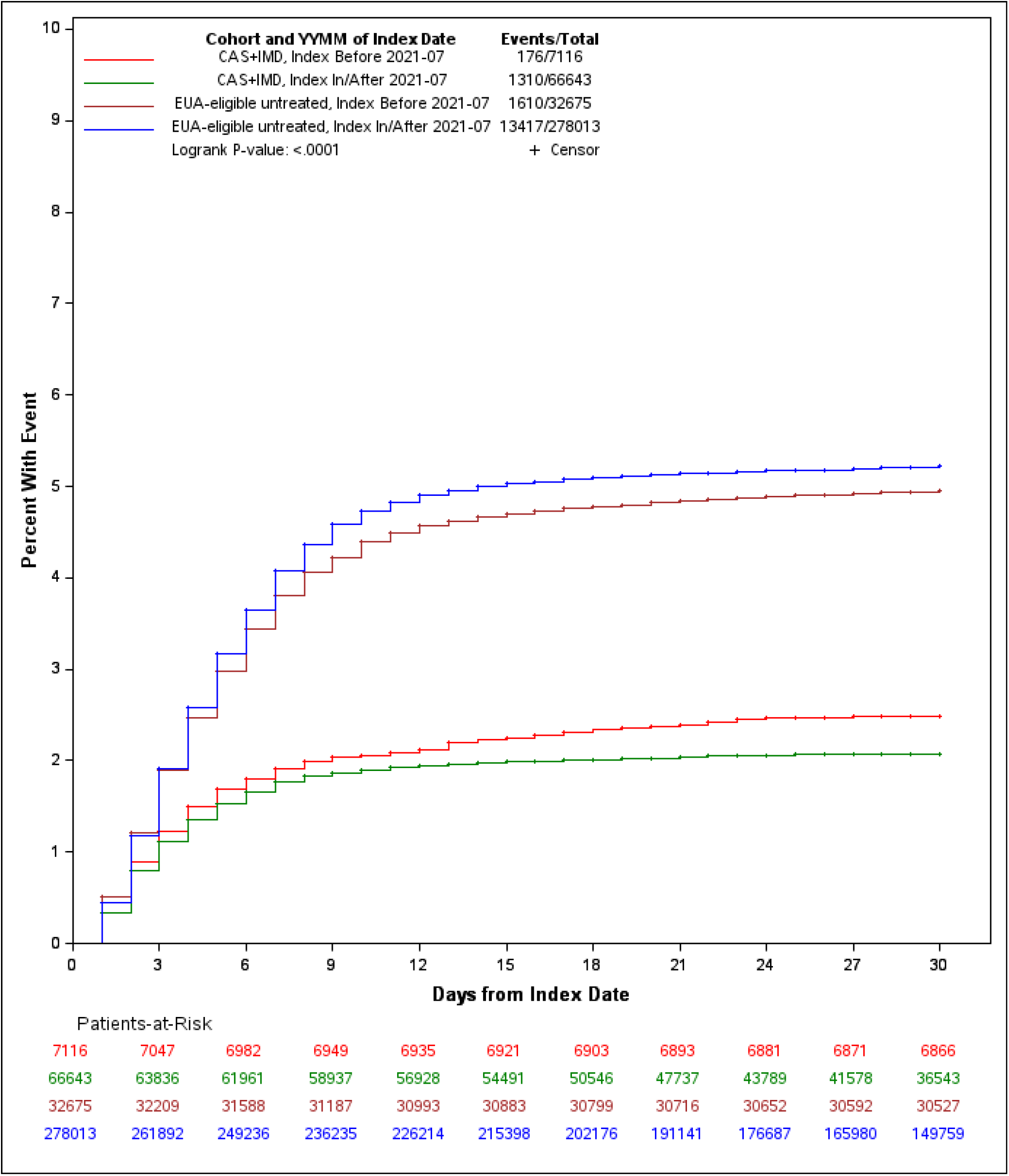
Kaplan-Meier Curve for Composite Endpoint of 30-day All-Cause mortality or COVID-19–Related Hospitalization Among Patients Diagnosed With COVID-19 in the Outpatient Setting, Stratified by Treatment Received and Timing of COVID-19 Diagnosis.

**Figure 4.**
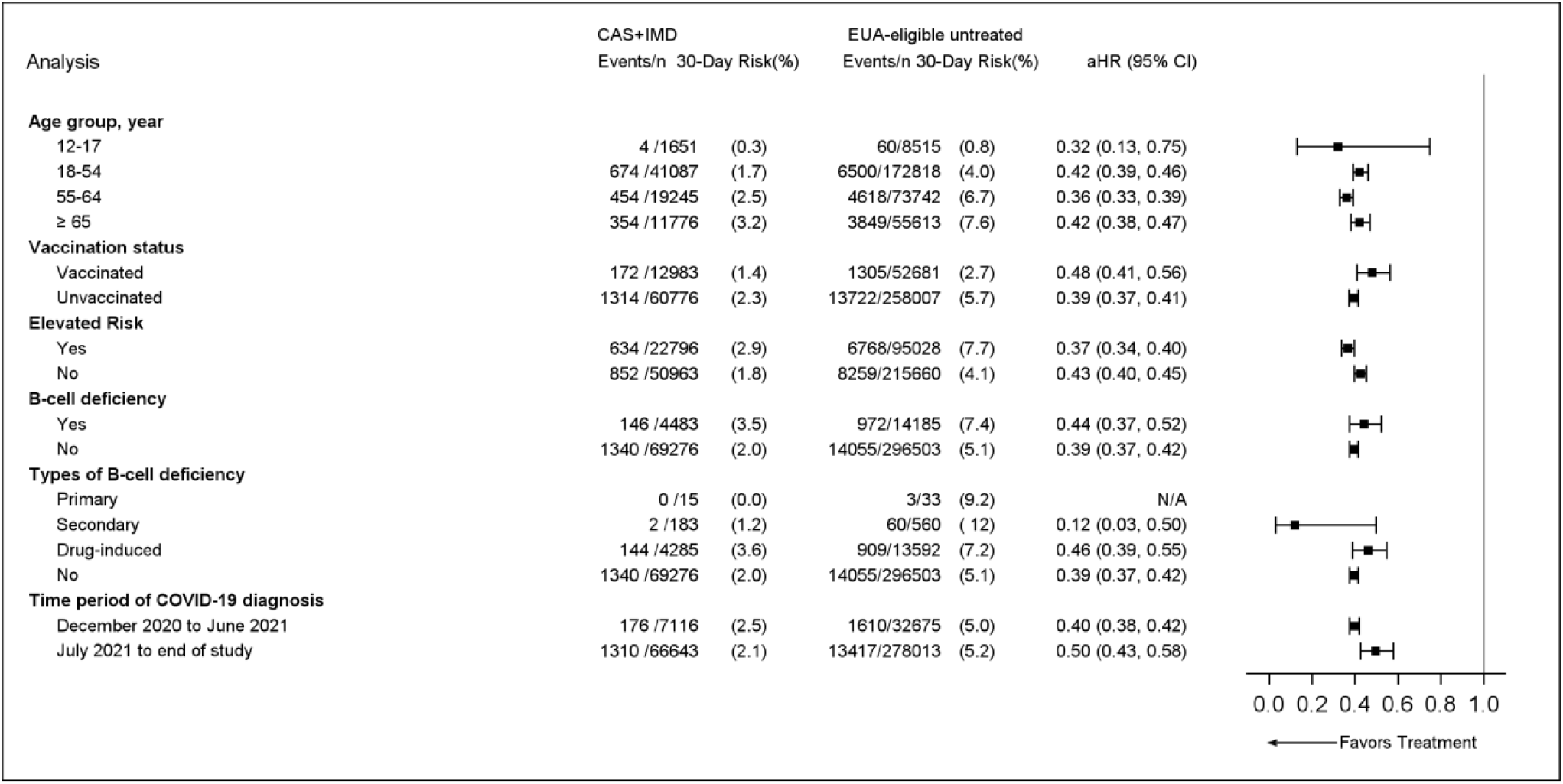
Subgroup Analyses of 30-Day Risk of All-Cause Mortality or COVID-19–Related Hospitalization Among Patients Diagnosed With COVID-19 in the Outpatient Setting. Abbreviations: aHR, adjusted hazard ratio; CI, confidence interval. ^a^ Defined as age ≥65 years or 55-64 years with BMI ≥35 kg/m^2^, type 2 diabetes, chronic obstructive pulmonary disease, or chronic kidney disease.

## DISCUSSION

This observational cohort study confirms and extends the evidence from clinical trials and other smaller real-world studies that patients with COVID-19 in the outpatient setting benefit from treatment with CAS+IMD. Among patients treated with CAS+IMD, there was a 60% reduction in the risk of 30-day all-cause mortality or COVID-19-related hospitalization compared to the EUA-eligible untreated patients. The benefit of treatment was observed across all patient subgroups. Notably, we found that the effectiveness of CAS+IMD was maintained during the Delta-dominant period and among patients receiving ≥1 dose of the COVID-19 vaccine.

In the primary analysis, the 60% reduction in risk is within the range of 50-90% lower risk of hospitalizations relative to untreated patients that was suggested by published real-world studies,^18,19,22,40,41^ although most of those studies used more broadly defined endpoints such as all-cause hospitalizations.^18,40,41^ Moreover, the risk reduction observed in this study is comparable to the 60% risk reduction of the same composite endpoint that was observed with CAS+IMD treatment compared to untreated EUA-eligible patients in a real-world study based on data from 2 large claims databases in the pre-Delta period.^42^ Our results are also consistent with those of the clinical trial demonstrating the efficacy of CAS+IMD for the treatment of COVID-19^2^ and suggest the benefits of CAS+IMD extend beyond the clinical trial setting. The subgroup analyses demonstrate that CAS+IMD is effective regardless of age or COVID-19 vaccination status, and is also effective in high-risk patient populations including those who are immunocompromised.

An important finding of this study was that the effectiveness of CAS+IMD was maintained when Delta was the dominant SARS-CoV-2 variant in the US. The Delta variant was of special concern because of its greater virulence in addition to its high transmissibility, which quickly made it the predominant variant.^8-12^ In the adjusted model, the aHRs showed that the magnitude of the treatment effect was slightly higher during the Delta-dominant period than the pre-Delta period (60% and 50% lower risk, respectively, relative to EUA-eligible untreated patients). While the observed difference may have been driven by lower accessibility and stricter patient selection in the pre-Delta period, our results demonstrate that CAS+IMD retains activity against the Delta variant. Such activity was previously suggested by *in vitro* studies^5,13-16^ and smaller real-world studies that reported its effectiveness for reducing all-cause hospitalization or mortality.^30,31^

An additional consequence of the emergence of variants such as Delta that are characterized by increased virulence is the occurrence of breakthrough infections among vaccinated individuals.^43-45^ Limited evidence has suggested that breakthrough infections in fully vaccinated patients are amenable to treatment with CAS+IMD.^19,31^ In the current analysis, the 52% reduction in risk among vaccinated patients suggests patients vaccinated against COVID-19 who experience breakthrough infections due to waning immunity or lack of effectiveness of vaccines can benefit from treatment.^19^ In addition, the reduction in outcome risk among patients treated with CAS+IMD was slightly greater (61%) among unvaccinated than vaccinated patients, likely resulting from a greater risk of severe COVID-19 among those who are not vaccinated. While these results support the benefits of treatment among those who cannot be or are unwilling to be vaccinated, they also suggest vaccinated individuals who contract COVID-19 and are eligible for treatment under the EUA can also benefit from treatment with CAS+IMD.

The findings of this study also suggest that immunocompromised patients (ie, B-cell deficient) can benefit from treatment with CAS+IMD. This is relevant because these patients have been shown to be at higher risk of being infected with COVID-19 and of progressing to more severe disease with poorer outcomes.^46,47^ Furthermore, patients with primary immunodeficiencies have a low likelihood of benefitting from vaccination,^48^ which makes them a group with a large unmet need.

### Limitations

Limitations of this study include that information on viral load and symptoms, variables that may predict severe COVID-19,^49-51^ are not captured in claims data. Moreover, a reason that at least some EUA-eligible patients were untreated may be that they had less severe disease, although social and cultural factors have also been reported to play a role in the decision for treatment with mAbs.^52^ If untreated patients had less severe disease than treated patients, the residual confounding would likely bias results against CAS+IMD. Another limitation is that several important variables such as BMI and COVID-19 vaccination status are not well captured in claims data; when this study was conducted, approximately 70% of the population had received 1 dose and 60% had received 2 doses. Residual confounding is likely, and if unvaccinated patients with higher BMIs are likely to have worse disease requiring treatment, it could result in underestimation of the effectiveness of CAS+IMD. We were also not able to distinguish between the subcutaneous and intravenous administration of CAS+IMD. Finally, the study period did not overlap with emergence of the Omicron variant, although CAS+IMD is not expected to be active against Omicron,^53^ as *in vitro* data indicate CAS+IMD has markedly reduced neutralization activity against this variant.^54,55^

## CONCLUSIONS

This study suggests that, in susceptible variants, treatment with CAS+IMD is effective against COVID-19 and its effectiveness is maintained across various patient subgroups. Among patients diagnosed with COVID-19 in the ambulatory setting, treatment with CAS+IMD was associated with a 60% reduction in risk of 30-day all-cause mortality or COVID-19-related hospitalization relative to matched untreated EUA-eligible patients that was maintained even after the emergence of the Delta variant and across a number of high-risk patient populations. While breakthrough infections are likely, especially in patients with risk factors and after emergence of VOC with reduced sensitivity to vaccines or with waning immunity, early treatment of these patients with CAS+IMD reduced the risk of disease progression that would require hospitalization or result in death. Evaluation of COVID-19 treatments and outcomes needs to remain ongoing as new VOC emerge so risk factors that can further improve COVID-19 management strategies can be identified.

## Supporting information

Hirshberg ICMJE Form

Murdock ICMJE Form

Weinreich ICMJE Form

Wang ICMJE Form

Jalbert ICMJE Form

Hussein ICMJE Form

Sanchez ICMJE Form

Mastey ICMJE Form

Wei ICMJE Form

## Data Availability

Qualified researchers may request access to study documents (including the clinical study report, study protocol with any amendments, blank case report form, and statistical analysis plan) that support the methods and findings reported in this manuscript.

## Author Contributions

All authors had full access to all of the data in the study and take responsibility for the integrity of the data and the accuracy of the data analysis.

*Concept and design:* Mohamed Hussein, Wenhui Wei, Vera Mastey, Robert J. Sanchez, Dana Murdock, David M. Weinreich, Jessica J. Jalbert

*Acquisition, analysis, or interpretation of data:* All authors.

*Drafting of the manuscript:* Mohamed Hussein, Wenhui Wei, Jessica J. Jalbert

*Critical revision of the manuscript for important intellectual content:* All authors.

*Statistical analysis:* Degang Wang, Wenhui Wei

*Administrative, technical, or material support:* Degang Wang

## Data Sharing

All data generated or analysed during this study are included in this published article.

## Additional Contributions

Medical writing support was provided by E. Jay Bienen, PhD, and was funded by Regeneron Pharmaceuticals, Inc. Data programming and analytics support was provided by Wenqin Qiang and Dehua Kan from KMK Consulting Inc. and was funded by Regeneron Pharmaceuticals, Inc.

