## Supplementary material for "Real-World Effectiveness of Casirivimab and Imdevimab Among Patients Diagnosed With COVID-19 in the Ambulatory Setting: A Retrospective Cohort Study Using a Large Claims Database"

### Supplementary Figure 1. Attrition of Study Cohorts

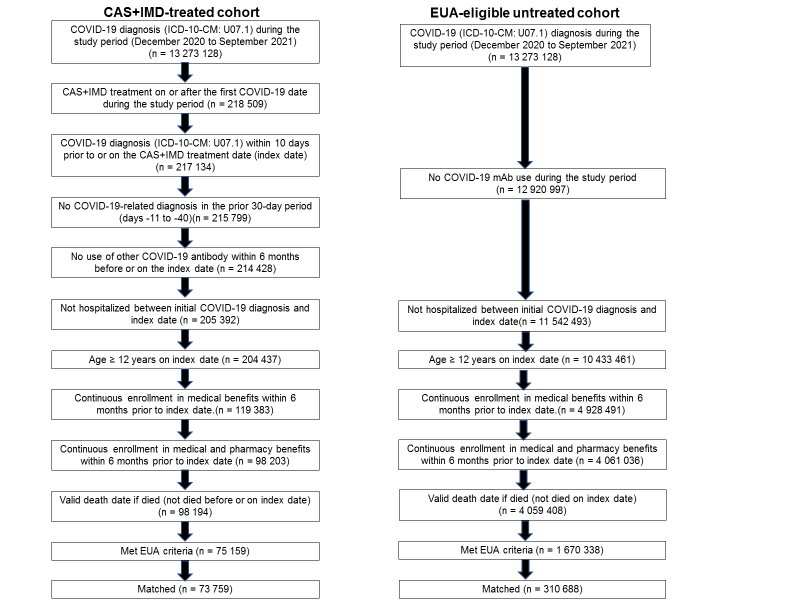

Abbreviations: CAS+IMD, casirivimab and imdevimab; EUA, Emergency Use Authorization.

### Supplementary Table 1. B-Cell Deficiencies

**Primary B-cell deficiencies (≥1 inpatient or ≥2 outpatient diagnoses ≥30 days apart during the baseline period, including index date)**

X-linked agammaglobulinemia

X-linked immunodeficiency with hyper-IgM

Selective IgA deficiency

Selective IgM deficiency

IgG subclass deficiency

Transient hypogammaglobulinemia of infancy

Common variable immunodeficiency (CVID)

Kappa/lambda light-chain deficiency

Severe combined immunodeficiency

Immunodysregulation polyendocrinopathyenteropathy X-linked syndrome (IPEX)

**Secondary causes of B-cell deficiency (≥1 inpatient or ≥2 outpatient diagnoses ≥30 days apart during the baseline period, including index date**

Multiple myeloma

Plasma cell leukemia

Acute lymphocytic leukemia (ALL)

Non-Hodgkin’s lymphoma (NHL)

Follicular lymphoma

Burkitt’s lymphoma

Diffuse B-cell lymphoma

Mantle cell lymphoma

Anaplastic large-cell lymphoma

Lymphoblastic lymphoma

Lymphoplasmacytic lymphoma

Marginal zone B-cell lymphoma/MALT lymphoma

Small-cell lymphocytic lymphoma

Non-Hodgkin’s lymphoma, unspecified/other

Hodgkin’s lymphoma (HL)

HIV/AIDS

**Drug-induced B-cell deficiencies (dispensation or administration within 3 months prior to index date)**

Rituximab

Ofatumumab (Kesimpta)

Ocrelizumab (Ocrevus)

Obinutuzumab

Inotuzumab ozogamicin

Blinatumomab

Alemtuzumab

Tocilizumab (Actemra)

Sarilumab (Kevzara)

Siltuximab

Belimumab (Benlysta)

Methotrexate

Mycophenolate mofetil (Cellcept, Myfortic)

Azathioprine (Imuran)

Systemic radiation therapy (excluding localized)

Chemotherapy

### Supplementary Table 2. Baseline Characteristics of the Unmatched Cohorts

| **Variable** | **CAS+IMD**  **(n = 75 159)** | **EUA-eligible untreated**  **(n = 1 670 338)** | **SMD^a^** |
| --- | --- | --- | --- |
| Age, years |  |  |  |
| Mean ± SD (range) | 49.9 ± 15.5 | 50.6 ± 20.3 | 0.04 |
| Median (IQR) | 51 (22) | 54 (30) | — |
| Range | 12-89 | 12-89 | — |
| Age group, years, n (%) |  |  |  |
| 12-17 | 1707 (2.3) | 158 360 (9.5) | 0.31 |
| 18-34 | 11 477 (15.3) | 246 752 (14.8) | 0.01 |
| 35-44 | 13 760 (18.3) | 199 898 (12.0) | 0.18 |
| 45-54 | 16 276 (21.7) | 233 978 (14.0) | 0.20 |
| 55-64 | 20 039 (26.7) | 384 835 (23.0) | 0.08 |
| 65-74 | 8240 (11.0) | 261 700 (15.7) | 0.14 |
| 75-84 | 2853 (3.8 | 120 818 (7.2) | 0.15 |
| ≥85 | 807 (1.1) | 63 997 (3.8) | 0.18 |
| Sex, n (%) |  |  |  |
| Female | 44 784 (59.6) | 1 001 478 (60.0) | 0.01 |
| Male | 30 375 (40.4) | 668 856 (40.0) | 0.01 |
| Region, n (%) |  |  |  |
| Midwest | 12 352 (16.4) | 322 581 (19.3) | 0.08 |
| Northeast | 7070 (9.4) | 359 449 (21.5) | 0.34 |
| South | 50 375 (67.0) | 768 800 (46.0) | 0.43 |
| West | 5362 (7.1) | 219 508 (13.1) | 0.20 |
| BMI category, n (%)^b^ |  |  |  |
| Not overweight | 2124 (2.8) | 59 172 (3.5) | 0.04 |
| Overweight (25-<30 kg/m^2^) | 6065 (8.1) | 114 849 (6.9) | 0.05 |
| Obese (30-<35 kg/m^2^) | 6932 (9.2) | 119 579 (7.2) | 0.08 |
| Severely obese (35-<40 kg/m^2^) | 5938 (7.9) | 122 395 (7.3) | 0.02 |
| Morbidly obese (≥40 kg/m^2^) | 8207 (10.9) | 146 253 (8.8) | 0.07 |
| Missing | 45 893 (61.1) | 1 108 090 (66.3) | 0.11 |
| CCI score, mean ± SD | 1.19 ± 1.75 | 1.37 ± 2.01 | 0.09 |
| Hospitalization during baseline period, n (%) | 10 396 (13.8) | 234 249 (14.0) | 0.01 |
| Month of index date, n (%) |  |  |  |
| December 2020 | 681 (0.9) | 380 085 (22.8) | 0.72 |
| January 2021 | 1933 (2.6) | 273 924 (16.4) | 0.49 |
| February 2021 | 1012 (1.4) | 115 495 (6.9) | 0.28 |
| March 2021 | 1058 (1.4) | 89 093 (5.3) | 0.22 |
| April 2021 | 1413 (1.9) | 80 755 (4.8) | 0.17 |
| May 2021 | 778 (1.0) | 44 338 (2.7) | 0.12 |
| June 2021 | 563 (0.8) | 23 354 (1.4) | 0.06 |
| July 2021 | 6598 (8.8) | 115 920 (6.9) | 0.07 |
| August 2021 | 31 584 (42.0) | 310 591 (18.6) | 0.53 |
| September 2021 | 29 539 (39.3) | 236 783 (14.2) | 0.59 |
| Time from diagnosis to index date, days |  |  |  |
| Mean ± SD | 1.6 ± 2.1 | 1.4 ± 2.0 | 0.11 |
| Median (IQR) | 1 (3) | 0 (2) | — |
| Range | 0-10 | 0-10 | — |
| Time from diagnosis to index date, days, n (%) |  |  |  |
| 0 | 33 993 (45.2) | 869 657 (52.1) | 0.14 |
| 1 | 12 837 (17.1) | 258 516 (15.5) | 0.04 |
| 2 | 9129 (12.2) | 180 320 (10.8) | 0.04 |
| ≥3 | 19 200 (25.6) | 361 845 (21.7) | 0.09 |
| Initial outpatient COVID-19 diagnosis in ER, n (%) | 22 486 (29.9) | 317 189 (19.0) | 0.26 |
| Vaccinated | 13 355 (17.8) | 150 812 (9.0) | 0.26 |
| EUA criteria |  |  |  |
| Age ≥65 years | 11 900 (15.8) | 446 515 (26.7) | 0.27 |
| Children overweight^c^ | 650 (0.9) | 55 980 (3.4) | 0.17 |
| Overweight | 27 142 (36.1) | 503 076 (30.1) | 0.13 |
| Pregnancy | 2665 (3.6) | 51 835 (3.1) | 0.03 |
| Chronic kidney disease | 3098 (4.1) | 110 048 (6.6) | 0.11 |
| Diabetes | 19 892 (26.5) | 482 885 (28.9) | 0.06 |
| Chronic pulmonary disease | 16 405 (21.8) | 325 594 (19.5) | 0.06 |
| Immunosuppressive disease | 8797 (11.7) | 194 593 (11.7) | 0.00 |
| Immunosuppressant use | 3860 (5.1) | 57 441 (3.4) | 0.08 |
| Sickle cell disease | 215 (0.3) | 3346 (0.2) | 0.02 |
| Cardiovascular disease, hypertension, or congenital heart disease | 41 316 (55.0) | 878 978 (52.6) | 0.05 |
| Neurodevelopmental disorders | 28 015 (37.3) | 687 561 (41.2) | 0.08 |
| Medical-related technological dependence | 19 400 (25.8) | 399 589 (23.9) | 0.04 |
| B-cell deficiency | 4975 (6.6) | 76 617 (4.6) | 0.09 |
| Primary | 19 (<0.1) | 221 (<0.1) | 0.01 |
| Secondary | 192 (0.3) | 3070 (0.2) | 0.02 |
| Drug-induced | 4764 6.3) | 73 326 (4.4) | 0.09 |
| Cancer or chemotherapy | 8838 (11.8) | 168 860 (10.1) | 0.05 |
| Cancer | 6359 (8.5) | 131 093 (7.9) | 0.02 |
| Chemotherapy | 3903 (5.2) | 59 532 (3.6) | 0.08 |

Abbreviations: BMI, body mass index; CAS+IMD, casirivimab and imdevimab; CCI, Charlson Comorbidity Index; ER, emergency room; EUA, Emergency Use Authorization; IQR, interquartile range; SD, standard deviation SMD, standardized mean difference.

^a^ Value ≥0.1 indicates significant imbalance between cohorts.

^b^ Based on diagnoses relating to the BMI categories.

^c^ Based on BMI ≥85th percentile for age and sex among those 12-17 years old.
